# Effective containment explains sub-exponential growth in confirmed cases of recent COVID-19 outbreak in Mainland China

**DOI:** 10.1101/2020.02.18.20024414

**Authors:** Benjamin F. Maier, Dirk Brockmann

## Abstract

The recent outbreak of COVID-19 in Mainland China is characterized by a distinctive algebraic, sub-exponential increase of confirmed cases with time during the early phase of the epidemic, contrasting an initial exponential growth expected for an unconstrained outbreak with sufficiently large reproduction rate. Although case counts vary significantly between affected provinces in Mainland China, the scaling law *t^μ^* is surprisingly universal, with a range of exponents *μ* = 2.1 ± 0.3. The universality of this behavior indicates that, in spite of social, regional, demographical, geographical, and socio-economical heterogeneities of affected Chinese provinces, this outbreak is dominated by fundamental mechanisms that are not captured by standard epidemiological models. We show that the observed scaling law is a direct consequence of containment policies that effectively deplete the susceptible population. To this end we introduce a parsimonious model that captures both, quarantine of symptomatic infected individuals as well as population wide isolation in response to mitigation policies or behavioral changes. For a wide range of parameters, the model reproduces the observed scaling law in confirmed cases and explains the observed exponents. Quantitative fits to empirical data permit the identification of peak times in the number of asymptomatic or oligo-symptomatic, unidentified infected individuals, as well as estimates of local variations in the basic reproduction number. The model implies that the observed scaling law in confirmed cases is a direct signature of effective contaiment strategies and/or systematic behavioral changes that affect a substantial fraction of the susceptible population. These insights may aid the implementation of containment strategies in potential export induced COVID-19 secondary outbreaks elsewhere or similar future outbreaks of other emergent infectious diseases.

## I. INTRODUCTION

The current outbreak of the new coronavirus in Mainland China (COVID-19, previously named 2019-nCoV) is closely monitored by governments, researchers, and the public alike [1–7]. The rapid increase of positively diagnosed cases in Mainland China and subsequent exportation and confirmation of cases in more than 20 countries worldwide raised concern on an international scale. The World Health Organization (WHO) therefore announced the COVID-19 outbreak a Public Health Emergency of International Concern [4].

Confirmed cases in Mainland China increased from approx. 330 on Jan. 21st, 2020 to more than 17,000 on Feb. 2nd, 2020 in a matter of two weeks [8]. In Hubei Province, the epicenter of the COVID-2019 outbreak, confirmed cases rose from 270 to 11,000 in this period, in all other Chinese provinces the cumulated case count increased from 60 to 6,000 in the same period.

An initial exponential growth of confirmed cases is generically expected for an uncontrolled outbreak and in most cases mitigated with a time delay by effective containment strategies and policies that reduce transmission and effective reproduction of the virus, commonly yielding a saturation in the cumulative case count and an exponential decay in the number of new infections. Although in Hubei the number of cases was observed to grow exponentially in early January [9], the subsequent rise followed a sub-exponential, super-linear, algebraic scaling law *t*^*μ*^ with an exponent *μ* = 2.3 (between Jan. 24th and Feb. 9th), cf. Fig. 1A. For the majority of the affected Chinese provinces of Mainland China, however, this type of algebraic rise occured from the beginning, lacking an initial exponential growth altogether. Surprisingly, the exponent *μ* does not vary substantially with a typical value of *μ* = 2.1 ± 0.3 for the confirmed case curves in other substantially affected provinces (confirmed case counts larger than 500 on Feb. 12th), despite geographical, socio-economical differences, differences in containment strategies, and heterogeneties that may have variable impacts on how the local epidemic unfolds, cf. Fig. 1B and C.

**Figure 1.**
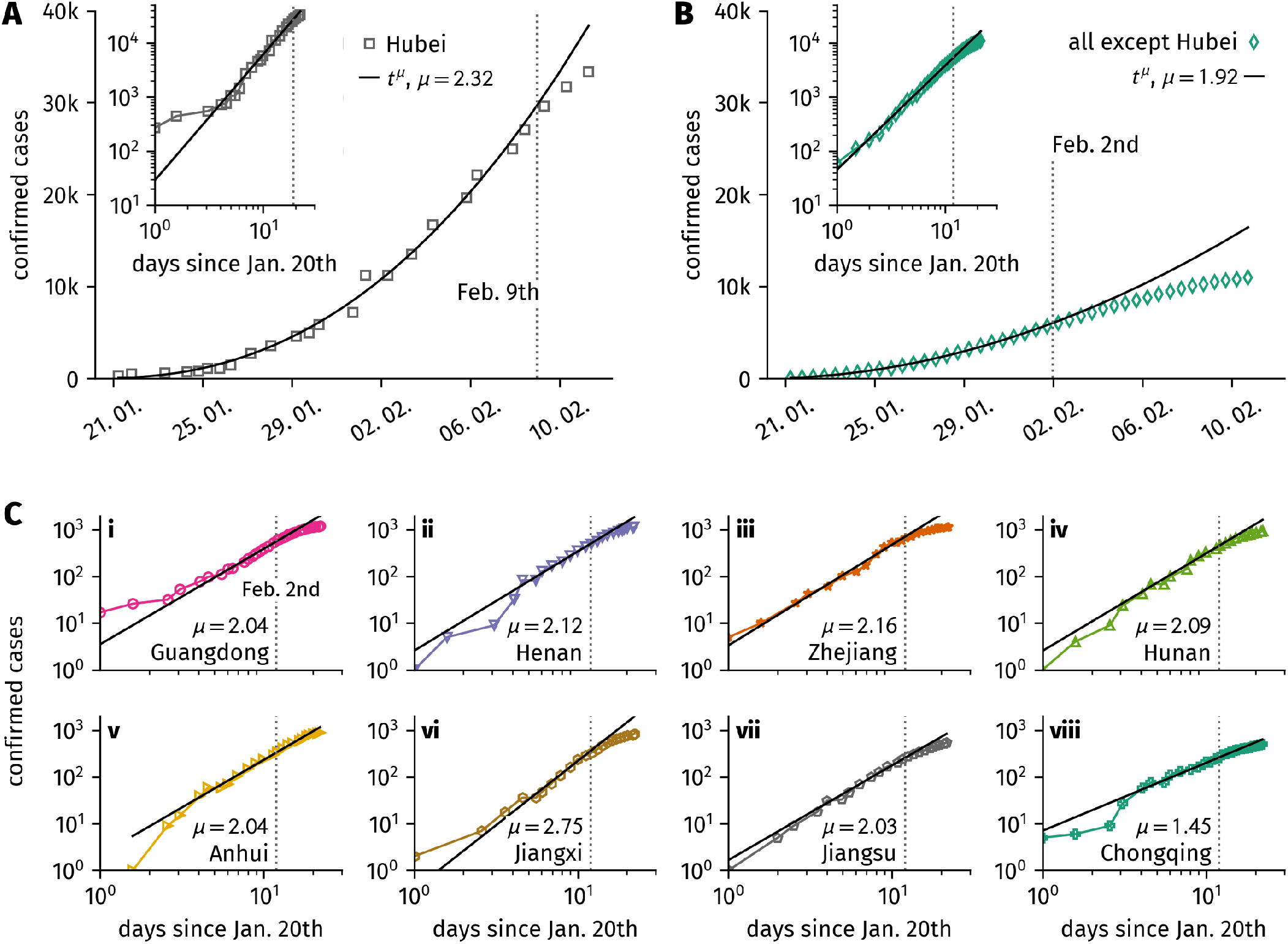
**A**: Confirmed case numbers in Hubei. The increase in cases follows a scaling law *t*^*μ*^ with an exponent *μ* = 2.32 after a short initial exponential growth phase. On Feb 9th the case count starts deviating towards lower values. **B**: Aggregated confirmed cases in all other affected provinces except Hubei. *C*(*t*) follows a scaling law with exponent *μ* = 1.92 until Feb. 2nd when case counts deviate to lower values. The insets in **A** and **B** depict *C*(*t*) on a log-log scale. **C**: Confirmed cases as a function of time for the 8 most affected provinces in China. The curves follow a scaling law with exponents *μ* ≈ 2 with the exception of Chongqing Province (*μ* = 1.45) and Jiangxi Province (*μ* = 2.75).

The universality of the scaling relation *t*^*μ*^ with similar exponents *μ* is evidence that this aspect of the dynamics is determined by fundamental principles that are at work and robust with respect to variation of other parameters that typically shape the temporal evolution of epidemic processes. Three questions immediately arise, (i) what may be the reason for this functional dependency, (ii) are provinces other than Hubei mostly driven by export cases from Hubei and therefore follow a similar functional form in case counts as suggested by preliminary studies [10–12], or, alternatively, (iii) is the scaling law a consequence of endogeneous and basic epidemiological processes and a consequence of a balance between transmission events and containment factors.

Here we propose a parsimonious epidemiological model that incorporates quarantine measures and containment policies. In addition to standard epidemiological parameters such as transmission and recovery rate, the model introduces effective quarantine measures that act on *both*, symptomatic infected individuals as well as susceptible individuals. We show that depleting both “ends” of the transmission interactions that involve an infectious and a susceptible individual naturally yields a sub-exponential, algebraic increase in confirmed cases. This behavior is qualitatively different from the behavior expected in a scenario in which only infecteds are targets of containment strategies to reduce transmission. In this case one expects either a stable exponential growth at a lower rate or an exponential decay if mitigation is sufficiently effective.

The model predicts the observed growth of case numbers exceptionally well for almost all affected provinces for epidemiological parameters estimated by preliminary studies [6, 7, 11, 13]. Furthermore, the model permits an indirect assessment of the peak time in the number of undetected infectious individuals (asymptomatic and oligosymptomatic cases) that preceeds the depletion of confirmed cases. This indicates that the number of unidentified infectious indviduals peaked in early February.

## II. SITUATION ASSESSMENT

### A. Case numbers in Hubei and other provinces

We rely on case number data provided by the Systems Science and Engineering group of Johns Hopkins University who currently give up to two updates per day on the number of laboratory-confirmed cases globally [8]. For the time period discussed in this paper, the group integrated and curated case data manually collected at four sources: the World Health Organization (WHO), the US Centers for Disease Control and Prevention (CDC), the European Centre for Disease Prevention and Control (ECDC), the Chinese physician’s platform DXY.cn, and the National Health Commission of the People’s Republic of China (NHC). The published data comprises total confirmed cases, total deaths, and total recovered cases as a function of time for each affected location. In the following we will focus on the number of total confirmed cases *C*(*t*).

In Hubei, we find that the initital increase in confirmed cases is followed by an algebraic scaling with time, i.e. *C*(*t*) ∝ *t*^*μ*^, with a scaling exponent *μ*≈ 2.3 that persisted until Feb. 9th, see Fig. 1A. On Feb. 12th, the case definition was changed by Chinese authorities which labeled a large number of previously unconfirmed cases as confirmed, leading to a discontinuity in the curves. We will therefore only consider data prior to Feb. 12th, 6am UTC.

Fig. 1B illustrates the cumulated case count in all affected provinces except Hubei province. In the period Jan. 21st until Feb. 2nd the curve follows an algebraic scaling law *t*^*μ*^ with *μ*≈1.9 lacking the initial exponential phase observed in Hubei province. Starting on Feb. 2nd, the observed case count starts deviating towards lower case counts. This poses the question whether the observed behavior is an averaging affect introduced by accumulating case counts across provinces. Interestingly, Fig. 1C provides evidence that this is not so. The confirmed case counts in the most affected provinces all exhibit a scaling law with exponents close to *μ* = 2. Among the most affected provinces only Chongqing Province exhibits a significantly lower exponent. Furthermore, all provincial case count curves start deviating from the algebraic curve around Feb. 2nd.

### B. Implemented policies

The Chinese government put several mitigation policies in place to contain the spread of the epidemic. In particular, positively diagnosed cases were either quarantined or put under a form of self-quarantine at home [14]. Similarly, suspicious cases were confined in monitored house arrest, e.g. individuals who arrived from Hubei before all traffic from its capital Wuhan was effectively restricted [15, 16]. These measures aimed at the removal of infectious individuals from the transmission process.

Additionally, measures were implemented that aimed at the protection of the susceptible population by the partial shutdown of public life. For instance, universities remained closed, many businesses closed down, and people were asked to remain in their homes for as much time as possible [14– 16]. These actions that affect both, susceptibles and non-symptomatic infectious individuals, not only protect susceptibles from acquiring the infection but also remove a substantial fraction of the entire pool of susceptibles from the transmission process and thus indirectly mitigate the proliferation of the virus in the population in much the same way that herd immunity is effective in the context of vaccine preventable diseases.

## III. EPIDEMIOLOGICAL MODELING WITH QUARANTINE AND ISOLATION

On a very basic level, an outbreak as the one in Hubei is captured by SIR dynamics [17]. The population is devided into three compartments that differentiate the state of invididuals with respect to the contagion process: (I)nfected, (S)usceptible to infection, and (R)emoved (i.e. not taking part in the transmission process). The corresponding variables *S, I*, and *R* quantify the respective compartments’ fraction of the total population such that *S* + *I* + *R* = 1. The temporal evolution of the number of cases is governed by two processes: The infection that describes the transmission from an infectious to a susceptible individual with basic reproduction number *R*_0_ and the recovery of an infected after an infectious period of average length *T*_*I*_. The basic reproduction number *R*_0_ captures the average number of secondary infections an infected will cause before he or she recovers or is effectively removed from the population.

Initially, a small fraction of infecteds yields an exponential growth if the basic reproduction number is larger than unity. When the supply of susceptibles is depleted, the epidemic reaches a maximum and the infecteds decline. A simple reduction of contacts caused by isolation policies could be associated with a reduction in the effective reproduction number, which would, however, still yield an exponential growth in the fraction of infecteds as long as *R*_0_ > 1, inconsistent with the observed scaling law *t*^*μ*^ discussed above. To test the hypothesis that the observed growth behavior can be caused by isolation policies that apply to both, infected and susceptible individuals, by effective public shutdown policies, we extend the SIR model by two additional mechanisms one of which can be interpreted as a process of removing susceptibles from the transmission process. First, we assume that general public containment policies or individual behavioral changes in response to the epidemic effectively remove individuals from the interaction dynamics or significantly reduce their participation in the transmission dynamics. Secondly, we account for the removal of symptomatic infected individuals by quarantine procedures. The dynamics is governed by the system of ordinary differential equations:

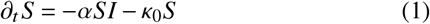

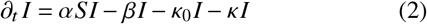

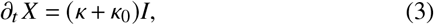

a generalization of the standard SIR model, henceforth referred to as the SIR-X model. The rate parameters *α* and *β* quantify the transmission rate and the recovery rate of the standard SIR model, respectively. Additionally, the impact of public containment is captured by the terms proportional to the containment rate κ_0_ that is effective in both *I* and *S* populations. Infected individuals are removed at rate κ corresponding to quarantine measures that only affect symptomatic infecteds. The new compartment *X* quantifies symptomatic, quarantined infecteds. Here we assume that the fraction *X t* is proportional to the empirical confirmed and reported cases. The case *κ*_0_ = 0 corresponds to a scenario in which the general population is unaffected by policies or does not commit behavioral changes in response to an epidemic. The case *κ* = 0 corresponds to a scenario in which symptomatic infecteds are not isolated specifically. Note that infecteds are always removed from their compartment *I* at a higher rate than susceptibles as *β* + *κ* + *κ*_0_ > *κ*_0_.

In the basic SIR model that captures unconstrained, free spread of the disease, the basic reproduction number *R*_0_ is related to transmission and recovery rate by *R*_0_ ≡ *R*_0,free_ = *α*/*β* because *β*^−1^ = *T*_*I*_ is the average time an infected individual remains infectious before recovery or removal. Here, the time period that an infected individual remains infectious is *T*_*I*,eff_ = (*β* + *κ*_0_ + *κ*)^−1^ such that the effective, or “observed” reproduction number *R*_0,eff_ = *αT*_*I*,eff_ is smaller than *R*_0,free_ since both *κ*_0_ > 0 and *κ* > 0.

We introduce *public containment leverage*

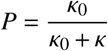

that reflects how strong isolation measures affect the general public in comparison to quarantine measures imposed on symptomatic infecteds alone. We further define

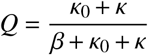

as the *quarantine probability*, i.e. the probability that an infected is identified and quarantined either in specialized hospital wards or at home.

The key mechanism at work in the model defined by Eqs. (1)-(3) is the exponentially fast depletion of susceptibles in addition to isolation of infecteds. This effect is sufficient to account for the observed scaling law in the number of confirmed cases for a plausible range of rate parameters as discussed below.

## IV. RESULTS

We assume that a small number of infected individuals travelled from Hubei to each of the other affected provinces before traffic restrictions were effective but at a time when containment measures were just being implemented. Fig. 2 illustrates the degree to which the case count for Hubei province and the aggregated case count for all other provinces is captured by the SIR-X model as defined by Eqs. (1)-(3).

**Figure 2.**
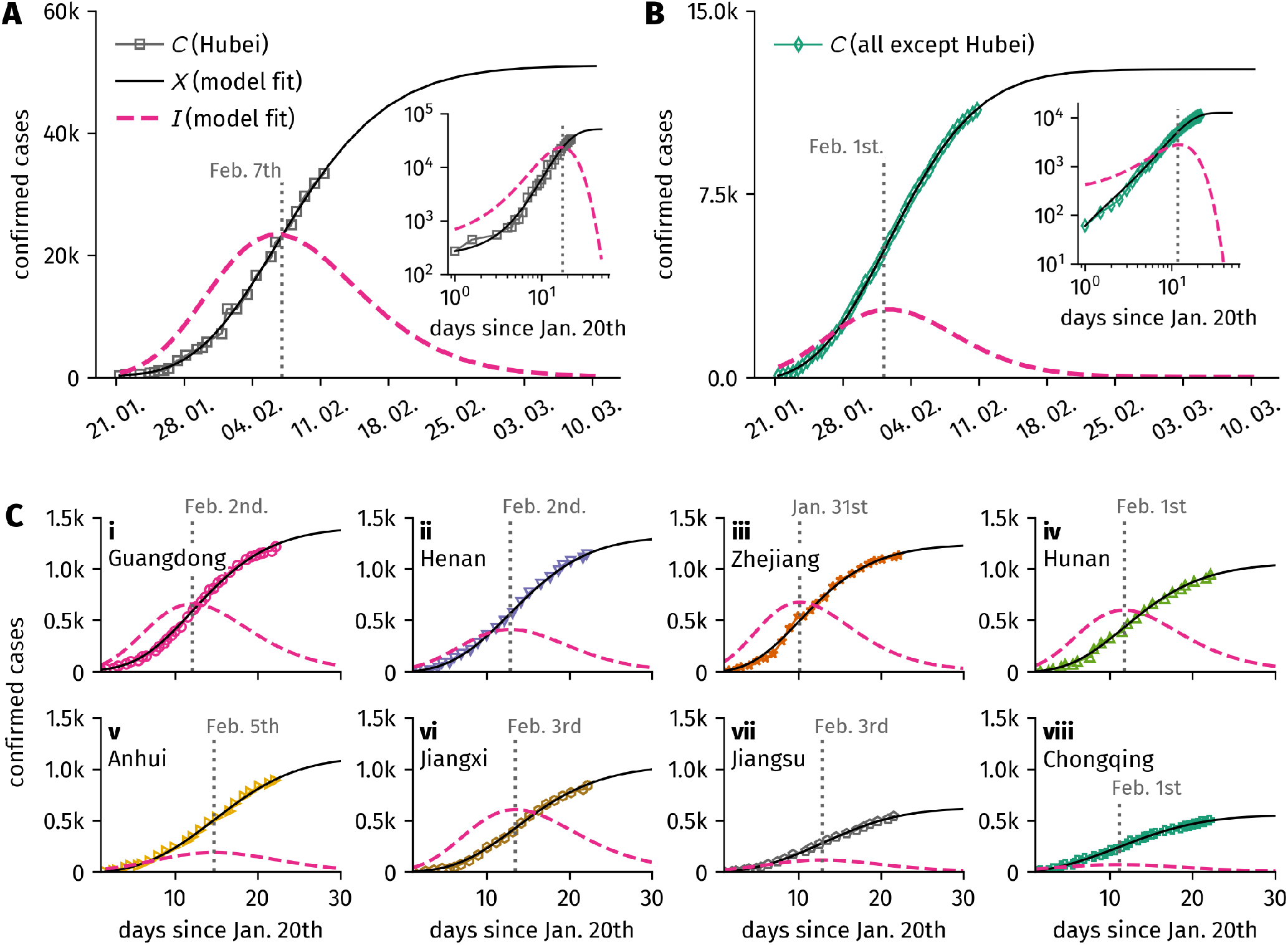
Case numbers in Hubei compared to model predictions. The quarantined compartment *X (t)* and the unidentified infectious compartment *I (t)* are obtained from least-squares fits of the model defined by Eqs. (1)-(3), cf. App. A. All fits were performed with fixed values of *R*_0,free_ = 6.2 and *T*_*I*_ = 8d. Note that the effective, observed basic reproduction number *R*_0,eff_ has lower values and varies for each of the affected provinces as discussed in Sec. IV and shown in Tab. I. **A:** In Hubei, the model captures both, the initial rise of confirmed cases as well as the subsequent algebraic growth. The confirmed cases are predicted to saturate at *C* = 51, 000. The model also predicts the time-course of the number of unidentified infectious individuals *I (t)* which peaks on Feb. 7th and declines exponentially afterwards. While the order of *I t* is associated with rather large fluctuations depending on the fitting procedure, the predicted peak time is robust, consistently around Feb. 7th. **B**: Model prediction for case numbers aggregated over all affected provinces other than Hubei. The case numbers’ algebraic growth is well reflected and predicted to saturate at *C* = 12, 600. In contrast to Hubei, the fraction of unidentified infecteds peaks around Feb. 1st, approximately a week earlier. The insets in **A** and **B** depict both data and fits on a log-log scale. **C**: Fits for confirmed cases as a function of time for the remaining 8 most affected provinces in China. All curves are well captured by the model fits that predict similar values for the peak time of unidentified infecteds.

We find that for a wide range of model parameters, the case count is well reproduced by the model. The model reproduces the scaling law *t*^*μ*^ as observed in the data for a significant period of time before saturating to a finite level. Remarkably, the model is able to reproduce both growth behaviors observed in the data: The model predicts the expected initial growth of case numbers in Hubei Province followed by an algebraic growth episode for ≈ 11 days until the saturation sets in, a consequence of the decay of unidentified infected individuals after a peak time around Feb. 7th (see Fig. 2A). Furthermore, the model also captures the immediate sub-exponential growth observed in the remaining most affected provinces (Fig. 2B-C). Again, saturation is induced by a decay of unidentified infecteds after peaks that occur several days before peak time in Hubei, ranging from Jan. 31st to Feb. 5th. For all provinces, following their respective peaks the number of unidentified infecteds (*I) t* decays over a time period that is longer than the reported estimation of maximum incubation period of 14 days [7, 13]. It is important to note that due to the uncertainty in the population size, the numerical value of unidentified infecteds is sensitive to parameter variations—the general shape of *I (t)*, however, is robust for a wide choice of parameters, as discussed in App. A.

Parameter choices for best fits were a fixed basic reproduction number of *R*_0,free_ = 6.2 (note that this reproduction number corresponds to an unconstrained epidemic) and a fixed mean infection duration of *T*_*I*_ = 8d consistent with previous reports concerning the incubation period of COVID-19 [7, 13]. The remaining fit parameters are shown in Tab. I. For these values, the effective basic reproduction number is found to range between 1.7 ≤ *R*_0,eff_ ≤ 3.3 for the discussed provinces, consistent with estimates found in previous early assessment studies [6, 7, 18, 19].

**Table I.** Fit parameters as described in Sec. III, fixed population size *N*, and resulting effective basic reproduction number *R*_0,eff_ for Hubei and the remaining majorly affected provinces discussed in the main text, decreasingly ordered by largest case number. The infectious period *T*_*I*_ and the basic reproduction number *R*_0,free_ = *αT*_*I*_ were fixed to values *T*_*I*_ = 8d and *R*_0,free_ = 6.2, respectively.

| Province | $N/10^6$ | $Q$ | $P$ | $I_0/X_0$ | $R_{0,\text{eff}}$ |
| --- | --- | --- | --- | --- | --- |
| Hubei | 57.1 | 0.47 | 0.66 | 2.55 | 3.28 |
| All exc. Hubei | 1277.8 | 0.69 | 0.21 | 6.98 | 1.93 |
| Guangdong | 104.3 | 0.51 | 0.75 | 3.66 | 3.02 |
| Henan | 94.3 | 0.61 | 0.38 | 41.00 | 2.41 |
| Zhejiang | 51.2 | 0.50 | 0.98 | 18.83 | 3.10 |
| Hunan | 67.0 | 0.47 | 1.00 | 58.34 | 3.28 |
| Anhui | 64.6 | 0.73 | 0.12 | 27.46 | 1.70 |
| Jiangxi | 44.0 | 0.44 | 1.00 | 19.41 | 3.46 |
| Jiangsu | 76.8 | 0.72 | 0.15 | 19.10 | 1.75 |
| Chongqing | 28.4 | 0.78 | 0.07 | 5.17 | 1.36 |

A detailed analysis of the obtained values for quarantine probability *Q* and public containment leverage *P* indicates that a wide range of these parameters can account for similar shapes of the respective case counts. Consequently, the model is structurally stable with respect to these parameters and the numerical value is of less importance than the quality of the mechanism they control (see App. A).

For the remaining 20 provinces, we find growth behaviors similar to the ones discussed before. Some follow a functional form comparable to the case counts in Hubei, others display a stronger agreement with pure algebraic growth, as can be seen in Fig. 3. Generally, a stronger agreement with algebraic growth can be associated with stronger public containment leverages.

**Figure 3.**
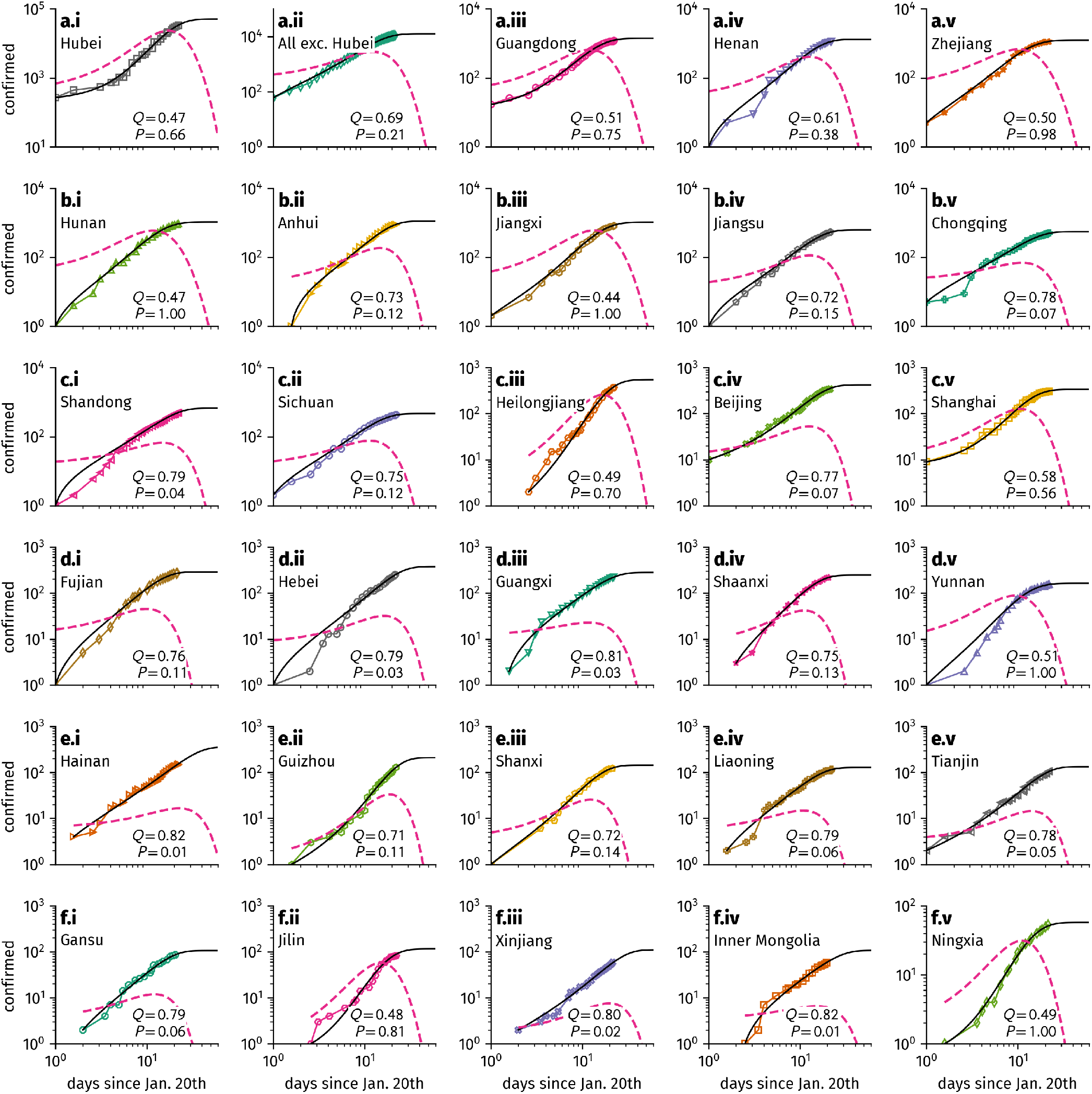
Case numbers of all affected provinces reproduced by the SIR-X model defined by Eqs. (1)-(3) and the fraction of unidentified infecteds *I (t)* as obtained from least-squares fits. All fits were performed with fixed values of *R*_0,free_ = 6.2 and *T*_*I*_ = 8d. For more details concerning the fitting procedure, cf. App. A.

In order to stress the general importance of public containment measures and to clarify to what extent quarantine is connected to case confirmation we also analyzed a model variant where (i) the general public and infecteds are removed from the transmission process in the same way and (ii) case counting is decoupled from the quarantine process (see App. C). We find that these model variants describe the real case count data reasonably well for the majority of provinces, further evidence of the importance of containment policies that target the susceptible population. In App. B we present further analytical evidence for this conclusion.

Note that the described saturation behavior of confirmed cases requires that eventually all susceptibles will effectively be removed from the transmission process. In reality, not every susceptible person can be isolated for such an extended period of time as the model suggests. One might expect instead that the number of infecteds will decay more slowly and saturate to a small, yet non-zero level. At this point it would be crucial to identify unquarantined infecteds more efficiently in order to completely shut down the transmission process. Due to potential difficulties in upholding the containment policies for such an extended period of time, we expect that our predictions will underestimate the final total amount of confirmed cases.

## V. DISCUSSION AND CONCLUSION

In summary, we find that one of the key features of the dynamics of the COVID-19 epidemic in Hubei Province but also in all other provinces is the robust sub-exponential rise in the number of confirmed cases according to the scaling law *t*^*μ*^ during the first episode of the epidemic. This general, almost universal behavior is evidence that fundamental principles are at work associated with this particular epidemic that are dominated by the interplay of the contagion process with endogeneous behavioral changes in the susceptible population and external mitigation policies. We find that the generic scaling of case counts with time is independent of many other factors that often shape the time-course of an epidemic.

The model defined by Eqs. (1)-(3) and discussed here indicates that this type of behavior can generally be expected if the supply of susceptible individuals is systematically decreased by means of implemented containment strategies or behavioral changes in response to information about the ongoing epidemic. Unlike contagion processes that develop without external interference at all or processes that merely lead to parametric changes in the dynamics, our analysis suggests that non-exponential growth is expected when the supply of susceptibles is depleted on a timescale comparable to the infectious period assiociated with the infection. The effective depletion of the susceptible populations on a timescale 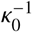 in the model does not imply that the pool of susceptibles must be depleted in a physical sense. Rather, this depletion could be achieved indirectly.

Containment that targets the susceptible population is a requirement for the observed algebraic scaling of case counts with time, unlike the reduction of transmission rates, or the reduction of the duration infected individuals remain infectious. Both of the latter measures address infected individuals only, which leads to a reduction of the exponential growth rate of new infections, or an exponential decay if the corresponding measures are sufficiently effective, but do not alter the functional growth behavior.

The fact that scaling laws were observed in Hubei after an initial exponential phase and that an exponential phase was not observed in the other substantially affected provinces is consistent with the model predictions if one assumes that the response in the population and the effect of policies lagged the onset of the epidemic in Hubei province and was immediate in all other provinces that were in an alert status already when cases were increasing substantially in Hubei province.

The model reproduces the empirical case counts in all provinces well for plausible parameter values. The quality of the reproduction of the case counts in all 29 affected provinces can be used to estimate the peak time of the number of asymptomatic or oligo-symptomatic infected individuals in the population, which is the key quantity for estimating the time when this outbreak will wane. The current analysis indicates that this peak time was reached around Feb. 7th for Hubei and within the first days of February in the remaining affected provinces.

The model further suggests that the public response to the epidemic and the containment measures put in place by the Chinese administration were effective despite the increase in confirmed cases. That this behavior was universally observed in all provinces also indicates that mitigation strategies were universally effective. Based on our analysis, we argue that the implemented containment strategies should stay in effect for a longer time than the incubation period after the saturation in confirmed cases sets in for this particular outbreak.

Our analysis further implies and provides evidence that mitigation strategies that target the susceptible population and induce behavioral changes at this “end” of the transmission process are most effective to contain an epidemic—especially in situations when asymptomatic or mildly symptomatic infectious periods are long or their duration unknown. This may be of importance for developing containment strategies in future scenarios or if the current COVID-19 epidemic were to trigger large scale outbreaks in other regions of the world by exports and subsequent proliferation.

## Data Availability

All data and analysis material is available in a separate GitHub repository.

https://github.com/benmaier/COVID19CaseNumberModel

## ACKNOWLEDGMENTS

We want to express our gratitude to L. H. Wieler and L. Schaade for helpful comments regarding the manuscript. B. F. M. is financially supported as an *Add-on Fellow for Interdisciplinary Life Science* by the Joachim Herz Stiftung.

## Appendix A: Fitting routine and analysis

Fits were performed using the Levenberg-Marquardt method of least squares. We fixed the epidemiological parameters to duration of infection *T*_*I*_ = 8d and basic reproduction number *R*_0,free_ = *αT*_*I*_ = 6.2. The population size *N* of each of the affected Chinese provinces was obtained from the Geonames project [20] and is listed in Tab. I. For each confirmed cases data set, we set the initial conditions *X (t*_0)_ = *C (t*_0)_ *N, I (t*_0)_ = (*I*_0_/*X*_0_) *X(t*_0_), and *S (t*_0_) = 1 − *I (t*_0)_ − *X (t*_0_) where *t*_*i*_, *C (t*_*i*_) is the *i*-th pair of a province’s time and aggregated confirmed case number. Since the number of unidentified infectious is unknown per definition, the prefactor *I*_0_ *X*_0_≤ 1 was chosen as a fit parameter. The remaining fit parameters were quarantine rate *κ* > 0 and containment rate *κ*_0_ > 0. For the fit procedure, Eqs. (1)-(3) were integrated using the Dormand– Prince method which implements a fourth-order Runge–Kutta method with step-size control, yielding *I* (*t)* and *X* (*t)* for every parameter configuration and every data set. The residuals were computed as ℛ(*t*_*i*_) = *NX* (*t*_*i*_) − *C* (*t*_*i*_).

We find that the model accurately reflects both the sub-exponential growth as well as the saturating behavior observed in the data discussed in the main text, with the obtained fit parameters displayed in Tab. I. For the 9 majorly affected provinces and the aggregated data over all provinces except Hubei, the quarantine probability is found to have similar values of *Q* = 0.6 ± 0.1. The public containment leverage is fluctuating more strongly with values ranging between *P* = 0.07 and *P* = 1, where generally, lower values appear concurrently with a higher quarantine probability, which suggests that stronger quarantine implementation requires less public isolation. Yet, we advise not to overinterpret the values of these fit parameters, as many different parameter values generate similar developments of confirmed cases.

As we fixed the population size to be equal to the total population of the respective provinces, one might also ask how system size changes the results—for instance, if the outbreak is contained in a small region of a province, the effective population available to the transmission process will be substantially smaller. Therefore, we repeated the fit procedure with *N* as a free fit parameter to find that the form of *X (t)* is not altered substantially for different values of *N*, while *I (t)* can vary more strongly as reflected by a larger variation in *I*_0_. This result suggests that the estimation of the number of infecteds is associated with a larger uncertainty. The general shape of *I(t)*, however, remains stable such that an inference of the peak time of unidentified infecteds is reasonable. Furthermore, the model favours larger values of the containment rate *κ*_0_ for larger population sizes in order to reproduce emprical data. This is a reasonable because the number of available transmission pathways grows quadratically with the population size, making it easier for the disease to spread faster. In order to still observe sub-exponential growth, a large part of the susceptible population has to be removed quickly. Consistently, we find that a decay of susceptibles is necessary to reliably obtain the observed growth behavior (i.e. *P* > 0, *κ*_0_ > 0).

Concerning variation in the fixed model parameters, larger values of up to *R*_0,free_ = 12 yield results similar to the ones described above when adjusting the infectious period *T*_*I*_ to larger values, as well. For lower values of *R*_0,free_ < 6, the model fails to reproduce the scaling laws observed in the data. Similarly, the fit results are reasonably robust against variations of the duration of infection in a range of *T*_*I*_ ∈ [6 d, 20 d] with concurrent adjustment of *R*_0,free_.

Similar effects are found for the remaining 20 affected provinces for which model fits are displayed in Fig 3. with parameters given in Tab. II. In general, provinces with larger values of public containment leverage *P* exhibit a stronger agreement with the hypothesis of algebraic growth.

**Table II.** Fit parameters as described in Sec. III, fixed population size *N*, and resulting effective basic reproduction number *R*_0,eff_ for the remaining affected provinces, decreasingly ordered by largest case number. The infectious period *T*_*I*_ and the basic reproduction number *R*_0,free_ = *αT*_*I*_ were fixed to values *T*_*I*_ = 8d and *R*_0,free_ = 6.2, respectively.

| Province | $N/10^6$ | $Q$ | $P$ | $I_0/X_0$ | $R_{0,\text{eff}}$ |
| --- | --- | --- | --- | --- | --- |
| Shandong | 94.2 | 0.79 | 0.04 | 18.86 | 1.33 |
| Sichuan | 87.2 | 0.75 | 0.12 | 9.74 | 1.58 |
| Heilongjiang | 38.2 | 0.49 | 0.70 | 6.13 | 3.19 |
| Beijing | 14.9 | 0.77 | 0.07 | 1.50 | 1.42 |
| Shanghai | 22.3 | 0.58 | 0.56 | 1.99 | 2.58 |
| Fujian | 36.9 | 0.76 | 0.11 | 16.10 | 1.47 |
| Hebei | 69.9 | 0.79 | 0.03 | 9.41 | 1.27 |
| Guangxi | 48.2 | 0.81 | 0.03 | 6.83 | 1.15 |
| Shaanxi | 37.6 | 0.75 | 0.13 | 4.33 | 1.55 |
| Yunnan | 45.4 | 0.51 | 1.00 | 14.99 | 3.01 |
| Hainan | 9.3 | 0.82 | 0.01 | 1.77 | 1.12 |
| Guizhou | 37.9 | 0.71 | 0.11 | 2.23 | 1.81 |
| Shanxi | 34.1 | 0.72 | 0.14 | 4.93 | 1.71 |
| Liaoning | 43.1 | 0.79 | 0.06 | 3.51 | 1.28 |
| Tianjin | 14.0 | 0.78 | 0.05 | 1.96 | 1.37 |
| Gansu | 26.3 | 0.79 | 0.06 | 2.50 | 1.30 |
| Jilin | 27.3 | 0.48 | 0.81 | 3.79 | 3.22 |
| Xinjiang | 21.3 | 0.80 | 0.02 | 1.13 | 1.22 |
| Inner Mongolia | 24.3 | 0.82 | 0.01 | 4.11 | 1.11 |
| Ningxia | 6.2 | 0.49 | 1.00 | 3.99 | 3.16 |

All data and the analysis material is available online [21].

## Appendix B: Case-number development in early phase of the epidemic

In a typical outbreak scenario only few people are infected initially, such that *S (*0) = 1−ε and *I(*0) = ε with ϵ « 1, which implies that the depletion of susceptibles available to the transmission process will be dominated by shutdown policies. This assumption effectively linearizes Eq. (1), yielding the solution

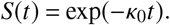

Further integration of the linearized Eq. (2) yields

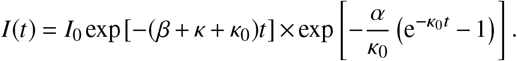

For small values of *t* we can expand the exponent to obtain the approximate growth function

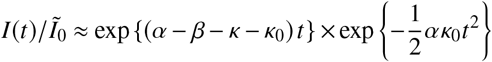

Here, the first factor implies that quarantining infecteds merely decreases the rate with which their number will grow exponentially, while the second term suppresses the whole transmission process well enough to alter the growth behavior. This implies that public shutdown policies facilitate epidemic containment in a more effective way than quarantine measures.

## Appendix C: Case confirmation without quarantine

In an alternative approach to the one discussed in the main text one may assume that isolation affects all citizens equally, i.e. *κ* = 0, but that the confirmation and counting of a new case is decoupled from whether the respective person is quarantined or not—rather, an infected is discovered and counted at rate 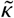. This implies an ODE system of

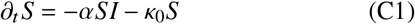

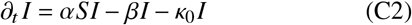

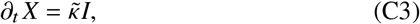

where as before, *X* will be equal to the number of confirmed cases *C*.

A numerical investigation of this alternative model shows that confirmed case data can be reasonably fit for all provinces, as well, which confirms the argumentation of App. B that removal of susceptibles from the transmission process is more effective than quarantine in mitigating epidemic spread.

